# Resting-state alterations in behavioral variant frontotemporal dementia are related to the distribution of monoamine and GABA neurotransmitter systems

**DOI:** 10.1101/2022.08.30.22278624

**Authors:** Lisa Hahn, Simon B. Eickhoff, Karsten Mueller, Leonhard Schilbach, Henryk Barthel, Klaus Fassbender, Klaus Fliessbach, Johannes Kornhuber, Johannes Prudlo, Matthis Synofzik, Jens Wiltfang, Janine Diehl-Schmid, Markus Otto, FTLD consortium, Juergen Dukart, Matthias L. Schroeter

## Abstract

Aside to clinical changes, behavioral variant frontotemporal dementia (bvFTD) is characterized by progressive structural and functional alterations in frontal and temporal regions. We examined if there is a selective vulnerability of specific neurotransmitter systems in bvFTD by evaluating the link between disease-related functional alterations and the spatial distribution of specific neurotransmitter systems and their underlying gene expression levels.

Maps of fractional amplitude of low frequency fluctuations (fALFF) were derived as a measure of local activity from resting-state functional magnetic resonance imaging for 52 bvFTD patients (mean age = 61.5 ± 10.0 years; 14 female) and 22 healthy controls (HC) (mean age = 63.6 ± 11.9 years; 13 female). We tested if alterations of fALFF in patients co-localize with the non-pathological distribution of specific neurotransmitter systems and their coding mRNA gene expression. Further, we evaluated if the strength of co-localization is associated with the observed clinical symptoms.

Patients displayed significantly reduced fALFF in fronto-temporal and fronto-parietal regions. These alterations co-localized with the distribution of serotonin (5-HT1b, 5-HT2a), dopamine (D2), and γ-aminobutyric acid (GABAa) receptors, the norepinephrine transporter (NET), and their encoding mRNA gene expression. The strength of co-localization with D2 and NET was associated with cognitive symptoms and disease severity of bvFTD.

Local brain functional activity reductions in bvFTD followed the distribution of specific neurotransmitter systems indicating a selective vulnerability. These findings provide novel insight into the disease mechanisms underlying functional alterations. Our data-driven method opens the road to generate new hypotheses for pharmacological interventions in neurodegenerative diseases even beyond bvFTD.

## Introduction

Frontotemporal lobar degeneration is the second most common type of early-onset dementia under the age of 65 years.[1] Its most common subtype, behavioral variant frontotemporal dementia (bvFTD), is characterized by detrimental changes in personality and behavior.[2] Patients can display both apathy and disinhibition, often combined with a lack of insight, and executive and socioemotional deficits.[3,4] Despite striking and early symptoms, bvFTD patients are often (i.e. up to 50%) misdiagnosed as having a psychiatric illness rather than a neurodegenerative disease.[5]

In addition to the presence of symptoms, the diagnosis requires consideration of family history due to its frequent heritable component and examination of different neuroimaging modalities.[2,6–8] Whereas atrophy in frontoinsular areas only occurs in later disease stages, glucose hypometabolism in frontal, anterior cingulate, and anterior temporal regions visible with fluorodeoxyglucose positron emission tomography (FDG-PET) is already detectable from an early stage onwards.[6,9] The fractional amplitude of low frequency fluctuations (fALFF) is a resting-state functional magnetic resonance imaging (rsfMRI) derived measure with good test-retest reliability that closely correlates with FDG-PET.[10,11] In frontotemporal dementia patients, fALFF was reduced in inferior parietal, frontal lobes and posterior cingulate cortex and holds great potential as MRI biomarker.[12,13] Low local fALFF activity in the left insula was linked to symptom deterioration.[14]

On a molecular level, frontotemporal lobar degeneration can be differentiated into three different subtypes based on abnormal protein deposition: tau (tau protein), transactive response DNA-binding protein with molecular weight 43 kDa (TDP-43), and FET (fused-in-sarcoma (FUS) and Ewing sarcoma (EWS) proteins, and TATA binding protein-associated factor 15 (TAF15)).[6,15] Whereas tau and TDP pathologies each occur in half of the bvFTD patients, FUS pathology is very rare.[16] Current research indicates prion-like propagation of these three proteins as disease mechanism.[17] Misfolded proteins accumulate and induce a self-perpetuating process so that protein aggregates can spread and amplify, leading to gradual dysfunction and eventually death of neurons and glial cells.[17] For example, tau can cause presynaptic dysfunction prior to loss of function or cell death,[18] whereas overexpression of TDP-43 leads to impairment of presynaptic integrity.[19] The role of FET proteins is not fully understood, although their involvement in gene expression suggests a mechanism of altered RNA processing.[20]

Neuronal connectivity plays a key role in the spread of pathology as it is thought to transmit along neural networks. Supporting the notion, previous studies also found an association between tau levels and functional connectivity in functionally connected brain regions, for example across normal ageing and Alzheimer’s disease.[21] Thereby, dopaminergic, serotonergic, glutamatergic, and GABAergic neurotransmission is affected. More specifically, current research indicates a deficit of neurons and receptors in these neurotransmitter systems.[17,22,23] Furthermore, these deficits have been associated with clinical symptoms. For example, whereas GABAergic deficits have been associated with disinhibition, increased dopaminergic neurotransmission and altered serotonergic modulation of dopaminergic neurotransmission have been associated with agitated and aggressive behavior.[24,25] Another study related apathy to glucose hypometabolism in the ventral tegmental area, a hub of the dopaminergic network.[3] Despite this compelling evidence of disease-related impairment at functional and molecular levels, the relationship between both remains poorly understood. It also remains unknown if the above neurotransmitter alterations reflect a disease-specific vulnerability of specific neuron populations or merely reflect a consequence of the ongoing neurodegeneration.

Based on the above findings, we hypothesize that the spatial distribution of fALFF and gray matter (GM) pathology in FTD will be related to the distribution of dopaminergic, serotonergic, and GABAergic neurotransmission. The aim of the current study was to gain novel insight into the disease mechanisms underlying functional and structural alterations in bvFTD by examining if there is a selective vulnerability of specific neurotransmitter systems. We evaluated the link between disease-related functional alterations and the spatial distribution of specific neurotransmitter systems and their underlying gene expression levels. In addition, we tested if these associations are linked to specific symptoms observed in this clinical population.

## Materials and Methods

### Participants

We included 52 patients with bvFTD (mean age = 61.5 ± 10.0 years; 14 female) and 22 age-matched healthy controls (HC) (mean age = 63.6 ± 11.9 years; 13 female) examined in nine centers of the German Consortium for Frontotemporal Lobar Degeneration (http://www.ftld.de) [26] into this study. Details regarding the distribution of demographic characteristics across centers are reported in Table S1. Diagnosis was based on established international diagnostic criteria.[27] Written informed consent was collected from each participant. The study was approved by the ethics committees of all universities involved in the German Consortium for Frontotemporal Lobar Degeneration, and was in accordance with the latest version of the Declaration of Helsinki. The clinical and neuropsychological test data included the Mini Mental State Exam (MMSE), Verbal Fluency (VF; animals), Boston Naming Test (BNT), Trail Making Test B (TMT-B), Apathy Evaluation Scale (AES) (companion-rated),[28] Frontal Systems Behavior Scale (FrSBe) (companion-rated) incl. subscales (executive function (EF), inhibition and apathy),[29] and Clinical Dementia Rating-Frontotemporal Lobar Degeneration scale-modified (CDR-FTLD).[30] Demographic and neuropsychological test information for both groups is displayed in Table 1.

**Table 1.**
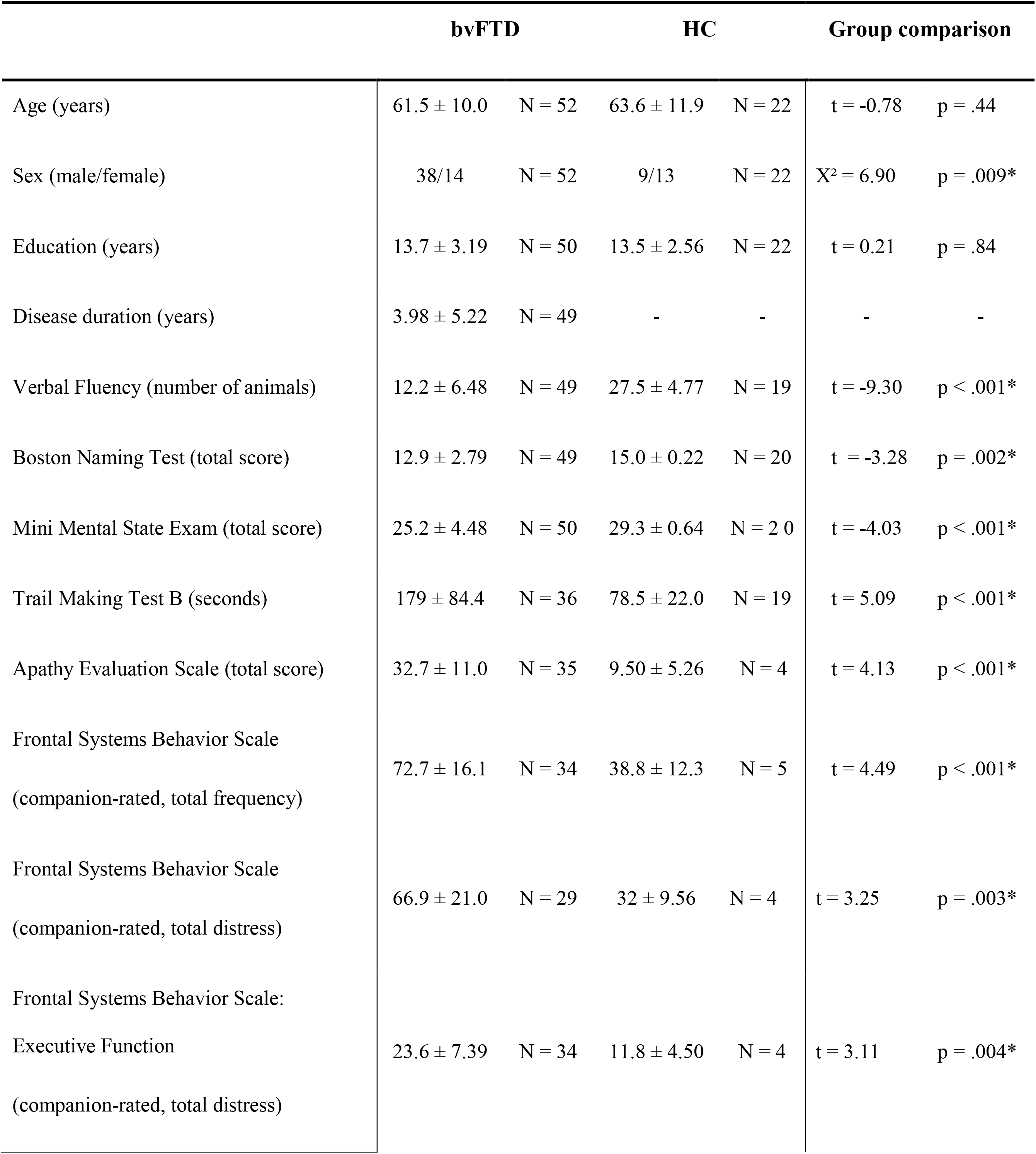
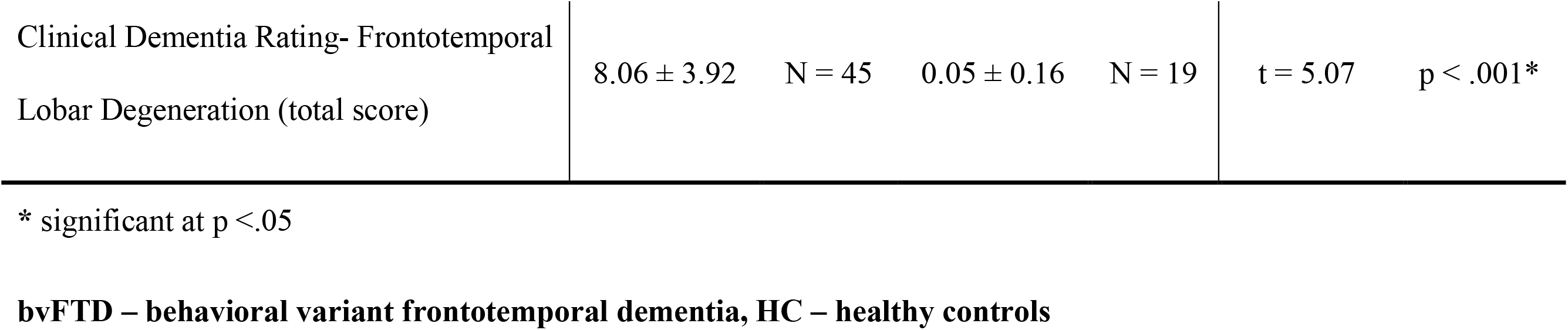
Demographic and clinical information for bvFTD patients and HC.

### MRI acquisition and preprocessing of imaging data

Structural T1-weighted magnetization-prepared rapid gradient-echo MRI and rsfMRI (TR = 2000 ms, TE = 30 ms, FOV = 64×64×30, voxel size = 3×3×5 mm, 300 volumes) were acquired on 3T devices. Table S2 reports center-specific imaging parameters confirming a high level of harmonization.

All initial pre-processing of imaging data was performed using SPM12.[31] To calculate voxel-wise gray matter volume (GMV), structural images were segmented, spatially normalized to MNI space, modulated, and smoothed by a Gaussian convolution kernel with 6 mm full-width at half maximum (FWHM). RsfMRI images were realigned, unwarped, co-registered to the structural image, spatially normalized to MNI space, and smoothed with a Gaussian convolution kernel with 6 mm FWHM. A gray matter mask was applied to reduce all analyses to gray matter tissue. Images were further processed in the REST toolbox [32] version 1.8. Mean white matter and cerebrospinal fluid signals as wells as 24 motion parameters (Friston-24) were regressed out before computing voxel-based measures of interest. fALFF was calculated at each voxel as the root mean square of the blood oxygen level dependent signal amplitude in the analysis frequency band (here: 0.01 – 0.08 Hz) divided by the amplitude in the entire frequency band.[32]

### Contrast analyses of fALFF and GMV

To test for fALFF alterations, group comparisons were performed in SPM12 using a flexible-factorial design with group (bvFTD or HC) as a factor and age, sex and site as covariates. To test for group differences in GMV, the same design with addition of total intracranial volume (TIV) was used. Pairwise group t-contrasts (i.e. HC>bvFTD, patients>bvFTD) were evaluated for significance using an exact permutation-based cluster threshold (1,000 permutations permuting group labels, p<.05) to control for multiple comparisons combined with an uncorrected voxel-threshold of p<.01.

### Spatial correlation with neurotransmitter density maps

Confounding effects of age, sex and site were regressed out from all images prior to further spatial correlation analyses. To test if fALFF alterations in bvFTD patients (relative to HC) are correlated with specific neurotransmitter systems, the JuSpace toolbox [33] was used. More specifically, we wanted to test if the spatial structure of fALFF maps in patients relative to HC is similar to the distribution of nuclear imaging derived neurotransmitter maps from independent healthy volunteer populations included in the toolbox (5-HT1a receptor, 5-HT1b receptor, 5-HT2a receptor, serotonin transporter (5-HTT), D1 receptor, D2 receptor, dopamine transporter (DAT), Fluorodopa (FDOPA), γ-aminobutyric acid type A (GABAa) receptors, μ-opioid (MU) receptors, and norepinephrine transporter (NET). Detailed information about the neurotransmitter maps is provided in Table S3. In contrast to standard analyses of fMRI data, this analysis might provide novel insight into potential neurophysiological mechanisms underlying the observed correlations.[33] Using the toolbox, mean values were extracted from both neurotransmitter and fALFF maps using gray matter regions from the Neuromorphometrics atlas. Extracted mean regional values of the patients’ fALFF maps were z-transformed relative to HC. Spearman correlation coefficients (Fisher’s Z transformed) were calculated between these z-transformed fALFF maps of the patients and the spatial distribution of the respective neurotransmitter maps. Exact permutation-based p-values as implemented in JuSpace (10,000 permutations randomly assigning group labels using orthogonal permutations) were computed to test if the distribution of the observed Fisher’s z-transformed individual correlation coefficients significantly deviated from zero. Furthermore, corrections for autocorrelations were performed. All analyses were false discovery rate (FDR) corrected for the number of tests (i.e. the number of neurotransmitter maps). Additionally, the receiver operating characteristic (ROC curves and corresponding areas under the curve (AUC) were calculated for patients (Fisher’s Z transformed Spearman correlations) vs. HC (leave-one-out Z-Score maps) to examine discriminability of the resulting fALFF-neurotransmitter correlations.

### Correlation with structural data

To test if the significant correlations observed between fALFF and neurotransmitter maps were driven by structural alterations (i.e. partial volume effects), the JuSpace analysis using the same parameters was repeated with local GMV incl. a correction for confounding effects of age, sex, site and TIV. For further exploration, fALFF and GMV Fisher’s Z transformed Spearman correlations as computed by the JuSpace toolbox were correlated with each other for each patient over all neurotransmitters. The median of those correlation coefficients was squared to calculate the variance in fALFF explained by GMV.

### Correlation with clinical data

To test if fALFF-neurotransmitter correlations are related to symptoms of bvFTD, we calculated Spearman correlation coefficients between significant fALFF-neurotransmitter correlations (Fisher’s z transformed Spearman correlation coefficients from JuSpace toolbox output, Fig. 2A) and clinical scales and neuropsychological test data (see Table 1). All analyses were FDR corrected for the number of tests.

### Association with gene expression profile maps

Furthermore, to test if fALFF alterations in bvFTD patients associated with specific neurotransmitter systems in the JuSpace analysis were also spatially correlated with their underlying mRNA gene expression profile maps, the MENGA toolbox [34] was used. Z-scores were calculated for the patients relative to HC using the confound-corrected images. The analyses were performed using 169 regions of interest and genes corresponding to each significantly associated neurotransmitter from the JuSpace analysis (5-HT1b: *HTR1B*; 5-HT2a: *HTR2A*; D2: *DRD2*; GABAa (nineteen subunits): *GABRA1-6, GABRB1-3, GABRG1-3, GABRR1-3, GABRD, GABRE, GABRP, GABRQ*; NET: *SLC6A2*). More specifically, Spearman correlation coefficients were calculated between the genomic values and re-sampled image values in the regions of interest for each patient and for each mRNA donor from the Allen Brain Atlas [35] separately. The Fisher’s Z transformed correlation coefficients were averaged over the six mRNA donors. Bonferroni-corrected one-sample t-tests were performed for each neurotransmitter to examine, whether the correlation coefficient differed significantly from zero.

### Neurotransmitter-genomic correlations and genomic autocorrelations

To further examine the association of fALFF-neurotransmitter correlations and mRNA gene expression profile maps, we explored the relationship between neurotransmitter maps included in the JuSpace toolbox and mRNA maps provided in the MENGA toolbox. The MENGA analysis was repeated using the same parameters to obtain Fisher’s Z transformed Spearman correlation coefficients between the neurotransmitter maps and the mRNA gene expression profile maps.

To evaluate the robustness of the mRNA maps between donors, genomic autocorrelations were calculated. Genomic autocorrelations were calculated per gene by obtaining Fisher’s Z transformed Spearman correlation coefficients between the genomic values of each of the six mRNA donors, which were then averaged.

## Results

### Contrast analysis of fALFF and GMV

First, we tested for group differences in fALFF between HC and patients. Compared to HC, bvFTD patients showed a significantly reduced fALFF signal in fronto-parietal and fronto-temporal regions (Fig. 1A). Furthermore, patients also showed reduced GMV in medial and lateral prefrontal, insular, temporal, anterior caudate and thalamic regions in comparison to HC (Fig. 1B). For a detailed representation of the thresholded fALFF and GMV T-maps see Fig. S1. Cluster size, incl. peak-level MNI coordinates and corresponding anatomical regions are reported in Table S4. For the distribution of Eigenvariates for the two groups in both modalities see Fig. S2.

**Fig. 1.**
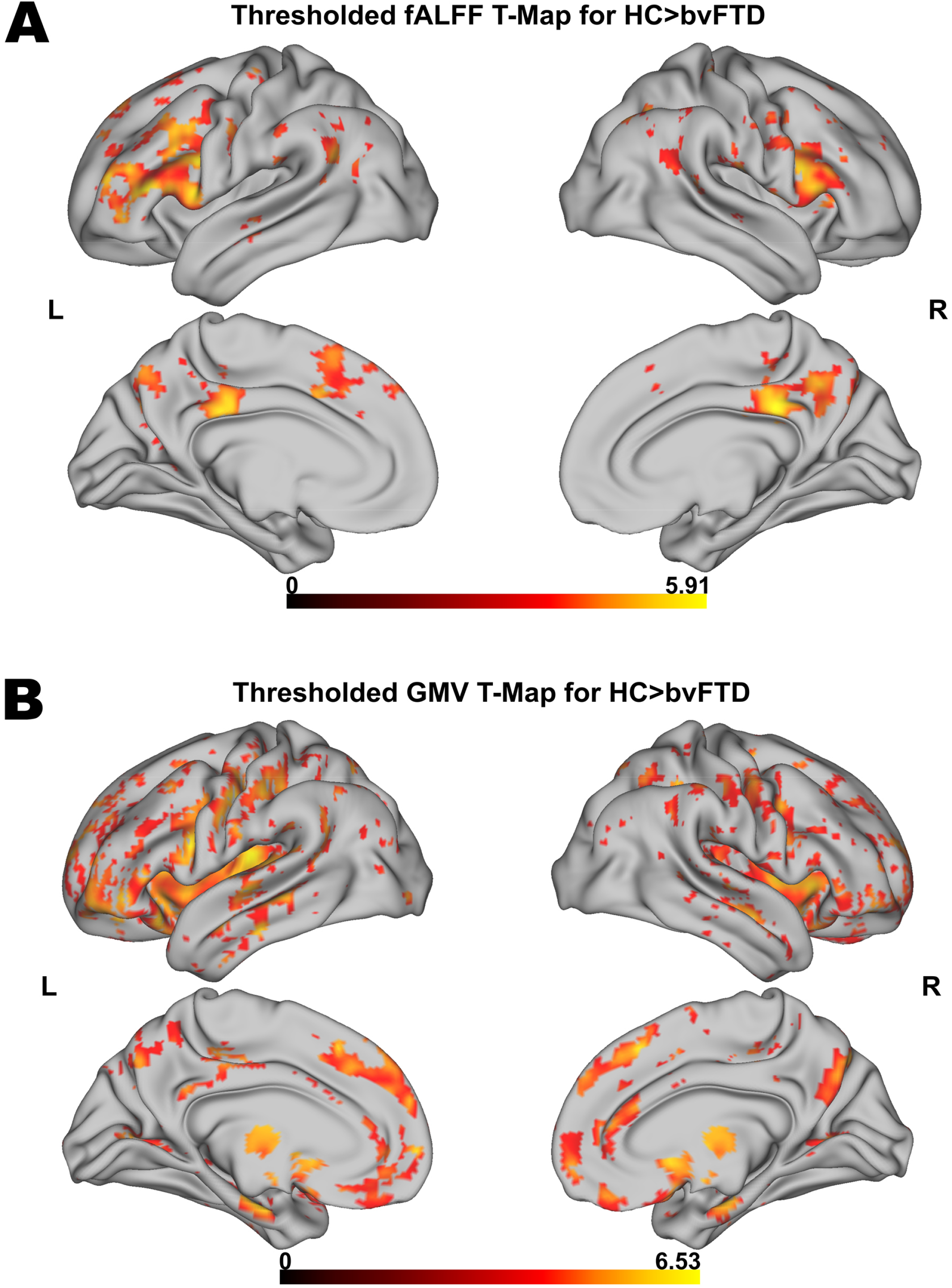
Voxel-wise results for fALFF and GMV group comparisons. Thresholded fALFF t-map **(A)** and thresholded GMV t-map **(B)** for HC>bvFTD using a permutation-based threshold (1000 permutations permuting group labels) at cluster-level p<.05 and voxel-level p<.001.

**Fig. 2.**
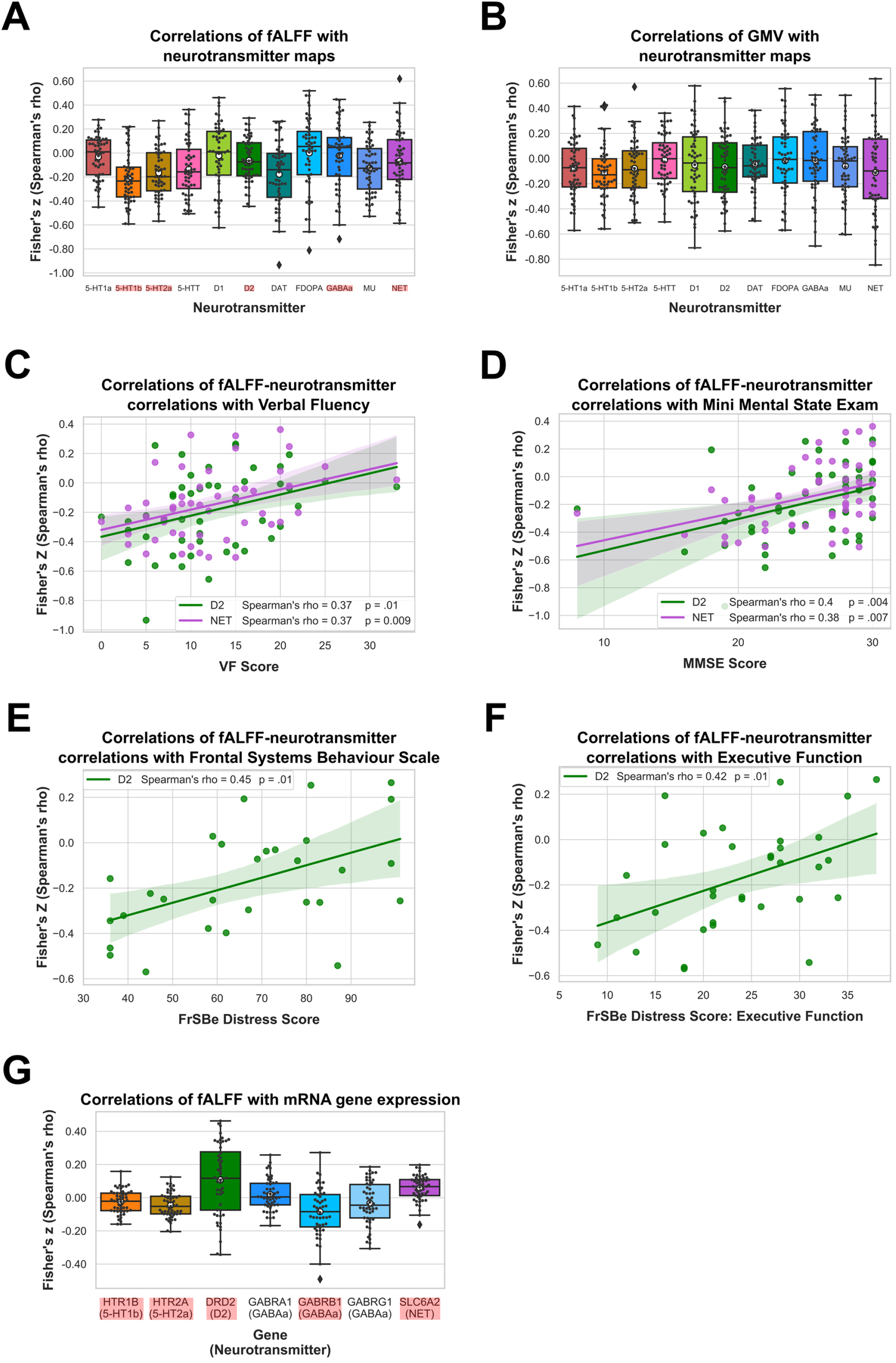
Results of spatial correlation analyses with in vivo and mRNA data. Correlation of fALFF **(A)** and GMV **(B)** with spatial distribution of neurotransmitter systems incl. 95% confidence intervals. Correlations of Verbal Fluency (N = 49) **(C)**, Mini Mental State Exam (N = 50) **(D)**, Frontal Systems Behavior Scale (N = 29) **(E)** and its Executive Function subscale (N = 34) **(F)** with fALFF-neurotransmitter strength of association incl. bootstrapped 95% confidence intervals. Correlations of fALFF with mRNA gene expression maps (N = 52) **(G)**. Statistically significant correlations in A, B, and G are marked in red and means are represented by white circles. Black circles in A, B and G represent individual Fisher’s z transformed Spearman correlation coefficients for each patient (N = 52) relative to controls with each neurotransmitter map. Colored circles in Figure C, D, E and F represent individual Fisher’s z transformed Spearman correlation coefficients between fALFF-neurotransmitter correlations from A and each neuropsychological scale.

### Spatial correlation with neurotransmitter maps

We performed correlation analyses to test if fALFF alterations in bvFTD significantly co-localize with the spatial distribution of specific neurotransmitter systems. fALFF alterations in bvFTD as compared to HC were significantly associated with the spatial distribution of 5-HT1b (mean *r* = -0.21, *p* < 0.001), 5-HT2a (mean *r* = -0.16, *p* = .0015), D2 (mean *r* = -0.18, *p* = .0057), GABAa (mean *r* = -0.12, *p* = .0157), and NET (mean *r* = -0.13, *p* = .0143) (*p*_*FDR*_ *=* .0157; Fig. 2A). The directionality of these findings (i.e. a negative correlation) suggest bvFTD displayed stronger reductions in fALFF relative to HC in areas which are associated with a higher non-pathological density of respective receptors and transporters. The AUC resulting from the ROC curves between Spearman correlation coefficients of patients and controls revealed a good discrimination for 5-HT1b (AUC = 0.74) and 5-HT2a (AUC = 0.71) and a fair discrimination for D2 (AUC = 0.69), GABAa (AUC = 0.68), and NET (AUC = 0.67) (Fig. 3A).

**Fig. 3.**
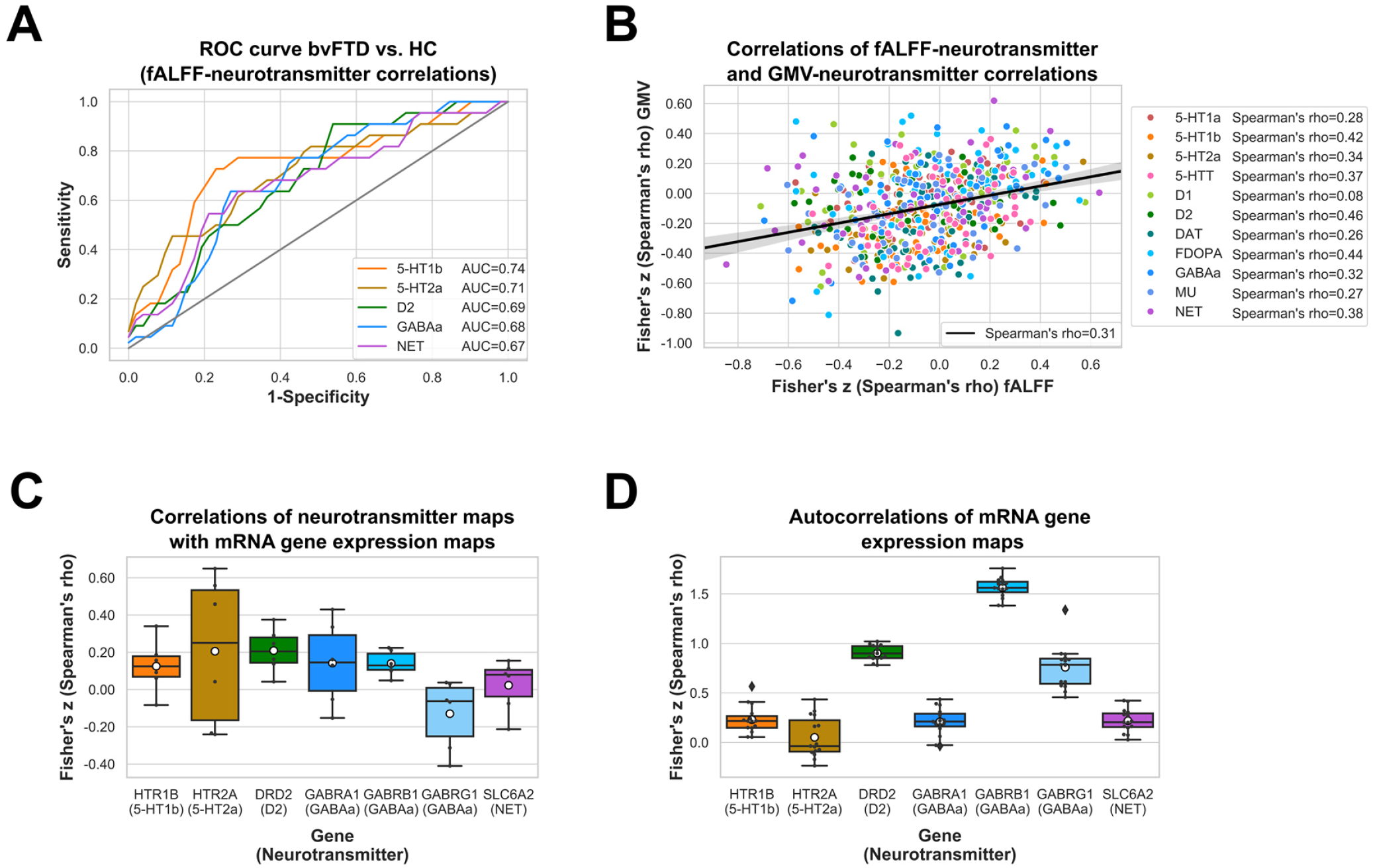
Results for fALFF-neurotransmitter ROC curve, correlations of fALFF-neurotransmitter and GMV-neurotransmitter correlations, correlations of neurotransmitter and mRNA gene expression maps, and autocorrelations of mRNA gene expression maps. Receiver operating characteristic (ROC) curves for HC vs. bvFTD are displayed for significant fALFF-neurotransmitter correlations (*N*_*bvFTD*_ = 52, *N*_*HC*_ = 22) **(A)**. Correlation of fALFF-neurotransmitter and GMV-neurotransmitter correlations are displayed for each patient and each significant neurotransmitter (N = 52) **(B)**. Correlations of neurotransmitter and mRNA gene expression maps **(C)** and autocorrelations of mRNA gene expression maps averaged across mRNA donors (N = 6) **(D)** are displayed for significant fALFF-neurotransmitter associations incl. 95% confidence intervals.

Next, we tested if similar co-localization patterns are observed with GMV. GMV alterations in bvFTD were not significantly associated with any of the neurotransmitter systems (Fig. 2B). fALFF-neurotransmitter and GMV-neurotransmitter correlations displayed a positive yet weak association with structural alterations explaining only 10% of variance in the fALFF alterations (Fig. 3B). All correlations and their corresponding permutation-based p-values are provided in Table S5. To exclude a potential bias caused by the collection of imaging data at different sites, we performed a Kruskal-Wallis Test to examine differences on the Fisher’s Z transformed correlations coefficients across sites. No significant differences (X^2^ = 6.34, *p* = .50, df = 7) were found among the sites.

### Relationship to clinical symptoms

Further, we tested if the significant fALFF-neurotransmitter correlation coefficients are also associated with symptoms or test results of bvFTD. The strength of fALFF co-localization with D2 and NET distribution was significantly associated with VF (D2: mean *r* = 0.37, *p* = .0092; NET: mean *r* = 0.37, *p* = .0086; N = 49; Fig. 2C) and MMSE (D2: mean *r* = 0.44, *p* = .0013; NET: mean *r* = 0.40, *p* = .0039; N = 50; Fig. 2D). Additionally, the strength of fALFF co-localization with D2 was significantly associated with the total distress score of FrSBe and its EF subscale (FrSBe: mean *r* = 0.45, *p* = .0145, N = 29; FrSBe-EF: mean *r* = 0.42, *p* = .0128, N = 34; Fig. 2E-F). The positive correlation coefficients suggest that more negative correlations between fALFF and neurotransmitter maps were associated with lower test performance, i.e. the higher/more fALFF reductions in areas with high neurotransmitter density, the lower the test performance. Associations with other neuropsychological tests were not significant (Table S6). We also tested if Eigenvariates extracted from the largest cluster of the HC>bvFTD contrast correlated with the specific symptoms of bvFTD (Table S7). None of the correlations remained significant after correction for multiple comparisons.

### Association with gene expression profile maps

Next, we evaluated if co-localization of fALFF is also observed with mRNA gene expression underlying the significantly associated neurotransmitter systems. For genes encoding the nineteen GABAa subunits, we first evaluated the variability between the subunits regarding their fALFF-mRNA correlations, their correlation with GABAa density and their mRNA autocorrelations (see Fig. S3). As the variability between the genes was high, we limited the analyses to genes encoding the three main subunits (GABRA1, GABRB1, GABRG1).

Correlations of fALFF alterations with mRNA gene expression profile maps in bvFTD relative to HC differed significantly from zero for *HTR1B* (encoding the 5-HT1b receptor; mean *r* = -0.02, *p* = .0144), *HTR2A* (encoding the 5-HT2a receptor; mean *r* = -0.04, *p* < .001), *DRD2* (encoding the D2 receptor; mean *r* = 0.11, *p* < .001), *GABRB1* (encoding subunit of the GABAa receptor; mean *r* = -0.08, *p* < .001) and *SLC6A2* (encoding NET; mean *r* = 0.06, *p* < .001), but not for GABRA1 (encoding subunit of the GABAa receptor; mean *r* = 0.02, *p* = .1414), GABRG1 (encoding subunit of the GABAa receptor; mean *r* = -0.03, *p* = .0730) (Fig. 2G). Thereby, correlations were negative for *HTR1B, HTR2A*, and *GABRB1*, i.e. fALFF was reduced in areas with higher expression of respective genes, and positive for *DRD2* and *SLC6A2*.

Further, we tested if there was an association between the neurotransmitter maps included in the JuSpace toolbox and the mRNA gene expression profile maps provided in the MENGA toolbox that were both derived from independent healthy volunteer populations. The correlations between spatial distributions of 5-HT1b, 5-HT2a, D2, GABAa, and NET, and corresponding mRNA gene expression profile maps were positive (5-HT1b/*HTR1B*: mean *r* = 0.12; 5-HT2a/*HTR2A*: mean *r* = 0.20; D2/DRD2: mean *r* = 0.21; GABAa/*GABRA1*): mean *r* = 0.14; GABAa/*GABRB1*: mean *r* = 0.14; NET/*SLC6A2*: mean *r* = 0.02) with exception of the GABRG1 gene (GABAa/*GABRG1*: mean *r* = -0.13)(Fig. 3C). Positive correlation coefficients suggest that higher neurotransmitter density was associated with higher expression of those neurotransmitters.

Lastly, to evaluate the robustness of the mRNA analyses, genomic autocorrelations were calculated. The genomic autocorrelation was high for *DRD2 (*mean *r* = 0.71), *GABRB1* (mean *r* = 0.92) and *GABRG1* (mean *r* = 0.64), small for *HTR1B* (mean *r* = 0.23), *SLC6A2* (mean *r* = 0.22) and *GABRA1* (mean *r* = 0.21), and very small for *HTR2A* (mean *r* = 0.05) (Fig. 3D).

## Discussion

In the current study, we examined if there is a selective vulnerability of specific neurotransmitter systems in bvFTD to gain novel insight into the disease mechanisms underlying functional and structural alterations. More specifically, we evaluated if fALFF alterations in bvFTD co-localize with specific neurotransmitter systems. We found a significant spatial co-localization between fALFF alterations in patients and the in vivo derived distribution of specific receptors and transporters covering serotonergic, dopaminergic, noradrenergic and GABAergic neurotransmission. These fALFF-neurotransmitter associations were also observed at the mRNA expression level and their strength correlated with specific clinical symptoms. All of the observed co-localizations with in vivo derived neurotransmitter estimates were negative with lower fALFF values in bvFTD being associated with a higher density of the respective receptors and transporters in health. The directionality of these findings supports the notion of higher vulnerability of respective networks to disease-related alterations. These findings are also largely in line with previous research concerning frontotemporal dementia (FTD) showing alterations in all of the respective neurotransmitter systems.[22,23]

The in vivo co-localization findings might also support the notion that prion-like propagation of proteins involved in bvFTD may align with specific neurotransmitter systems.[17] With regard to other brain disorders, linking functional connectivity with receptor density and expression, recent studies found an association between functional connectivity and receptor availability in schizophrenia, and an association between structural-functional decoupling and receptor gene expression in Parkinson’s disease.[36,37] A potential mechanism for the selective vulnerability of specific neurotransmitter systems is the propagation of proteins along functionally connected networks that has been previously demonstrated for various neurodegenerative diseases.[38,39] For example, in Alzheimer’s disease and normal ageing, tau levels closely correlated with functional connectivity.[21] We found moderate to large AUC when using the strength of the identified co-localizations for differentiation between patients and healthy controls suggesting that these findings may represent a measure of the affectedness of respective neurotransmitter systems. In bvFTD, neurodegeneration is thought to progress through the salience network involved in socioemotional tasks, which comprises the anterior cingulate and fronto-insular cortex, as well as the amygdala and the striatum.[6,17] The four neurotransmitter systems found to be deficient in our sample are relevant for the functioning of these structures (anterior cingulate cortex: e.g. serotonin, noradrenaline;[40,41] fronto-insular cortex: e.g. dopamine;[42] amygdala: e.g. GABA, dopamine, serotonin;[43] striatum: e.g. GABA, dopamine [44,45]). Although prion-like spread of misfolded proteins through the salience network provides a potential disease mechanism, further research of the exact mechanisms involved is needed.

For GMV, we did not find any significant co-localization with specific neurotransmitter systems. As the correlations with GMV showed a distinct pattern to fALFF and the variance explained by GMV in the observed fALFF-neurotransmitter associations was small, the observed associations with fALFF seem to be driven indeed by functional alterations and not by the underlying atrophy of respective regions. As prion-like propagation of misfolded proteins leads to a gradual dysfunction and eventually cell death,[17] some regions displaying high density of a specific neurotransmitter might suffer dysfunction (i.e. functional alterations), whereas others might already be exposed to cell death (i.e. structural alterations/atrophy).

The strength of co-localization of fALFF with D2 and NET was correlated with verbal fluency and dementia measures, both being impaired in patients with FTD.[46] Thereby, a stronger negative co-localization (i.e. lower fALFF in patients in high density regions in health) was associated with decreased test performance. These findings are in line with the reported role of dopamine in verbal fluency.[47] In contrast to the study by Murley et al.,[25] who reported an association of GABA deficits in FTD with disinhibition, we did not find this association. Beside the use of different methodology, a potential explanation may constitute the use of different inhibition measures. Whereas we measured disinhibition using the FrSBe, Murley et al. [25] used a stop-signal task.

Although, except for α1 and γ1 GABAa subunits, all of the co-localizations with fALFF identified with in vivo estimates were also significant at the respective mRNA gene expression level, we found correlation coefficients of both directionalities. Interestingly, whereas these correlations were solely negative for the in vivo derived maps, the correlations with gene expression profile maps were positive for D2 and NET, and negative for 5-HT1b, 5-HT2a and β1 GABAa subunit. Thus, for D2 and NET, we observed higher fALFF values in bvFTD patients in areas with high mRNA gene expression in health, whereas for 5-HT1b, 5-HT2a and β1 GABAa subunit we observed lower fALFF values in bvFTD patients in areas with high mRNA gene expression in health. One explanation for these seemingly contradictory findings is that mRNA gene expression seems to vary strongly between individuals. In our mRNA gene expression profile maps, the autocorrelation between mRNA donors was low for 5-HT1b, 5-HT2a, and α1 GABAa subunit, and NET, limiting the confidence in some of these findings. Additionally, the association of mRNA expression with protein products may also vary greatly between genes, being not associated at all or even negatively associated for some, and strongly correlated for others.[48,49] Potential reasons for the lack of or even negative correlations may be a decoupling in time as well as that other levels of regulation overrode the transcriptional level.[48] We observed a similar phenomenon in our data with the correlation of neurotransmitter density maps with their underlying mRNA gene expression being weak for all neurotransmitters except D2, β1 and γ1 GABAa subunits.

The current study was limited by the unavailability of medication information. Therefore, we were not able to control for its potential confounding effects. However, as bvFTD medication is typically restricted to serotonin reuptake inhibitors its effects should be primarily associated with availability of 5-HTT and directionally negate the effects of the disease. Furthermore, as the included PET maps were derived from healthy subjects, the applied approach only tests for co-localization of imaging changes with the non-pathological distribution of the respective neurotransmitter systems.

To summarize, we found fALFF reductions in bvFTD to co-localize with the in vivo and ex vivo derived distribution of serotonergic, dopaminergic, GABAergic, and noradrenergic neurotransmitter systems, pointing to a crucial vulnerability of these neurotransmitters. The strength of these associations was linked to some of the neuropsychological deficits observed in this disease. We propose a combination of spread of pathology through neuronal connectivity and more specifically, through the salience network, as a disease mechanism. Thereby, these findings provide novel insight into the mechanisms underlying the spatial constraints observed in progressive functional and structural alterations in bvFTD. Our data-driven method might even be used to generate new hypotheses for pharmacological intervention in neuropsychiatric diseases beyond this disorder.

## Data Availability

The original data supporting the findings of this study are available from the senior author (Matthias Schroeter) upon reasonable request. All derived statistical measures used here are available from the first author upon request. The software applied is publicly available at https://github.com/juryxy/JuSpace.

## Acknowledgements

Finally, we would like to acknowledge the Clinic for Cognitive Neurology in Leipzig, Annerose Engel, Anke Marschhauser, and Maryna Polyakova.

## Author contributions

LH and JD performed all analyses. KM, HB, KF, KF, JH, JP, MS, JW, JD, MO and MS were involved in the planning of the study and the data collection. All authors contributed to the final manuscript.

