## Supplementary Material for "Resting-state alterations in behavioral variant frontotemporal dementia are related to the distribution of monoamine and GABA neurotransmitter systems"

**Table S1** Distribution of number of subjects, age and sex across centers.

| Center | N | | Age (Mean ± Std.) | | Sex (male/female) | |
|---|---|---|---|---|---|---|
| | **HC** | **bvFTD** | **HC** | **bvFTD** | **HC** | **bvFTD** |
| Bonn | --- | 6 | --- | 66.7 ± 7.74 | --- | 6/0 |
| Erlangen | 1 | 4 | 74.0 ± 0.00 | 60.5 ± 4.65 | 1/0 | 3/1 |
| Göttingen | 1 | 5 | 37.0 ± 0.00 | 66.8 ± 7.43 | 0/1 | 5/0 |
| Homburg | 6 | 5 | 56.0 ± 10.5 | 59.8 ± 12.6 | 4/2 | 4/1 |
| Leipzig | 4 | 13 | 61.5 ± 8.23 | 61.8 ± 12.6 | 2/2 | 8/5 |
| München (TU) | --- | 13 | --- | 60.3 ± 8.87 | --- | 8/5 |
| Rostock | 8 | 4 | 73.4 ± 5.66 | 59.5 ± 9.54 | 2/6 | 2/2 |
| Tübingen | --- | 2 | --- | 48.5 ± 7.78 | --- | 2/0 |
| Ulm | 2 | --- | 59.5 ± 2.12 | --- | 0/2 | --- |

Bonn – University of Bonn, German Center for Neurodegenerative Diseases (DZNE), University Hospital Bonn
Erlangen – University Hospital Erlangen
Göttingen – Medical University Göttingen
Homburg – Saarland University Hospital
Leipzig – Max-Planck-Institute for Human Cognitive and Brain Sciences
TU München – Technical University of Munich
Rostock – University Hospital Rostock, German Center for Neurodegenerative Diseases (DZNE)
Tübingen – University Hospital Tübingen, Centre for Neurology, Hertie-Institute for Clinical Brain Research
Ulm – Ulm University

**Table S2** Center-specific imaging parameters for structural and functional imaging.

| Center | rsfMRI | | | | | Structural MRI | | | |
|---|---|---|---|---|---|---|---|---|---|
| | TE (ms) | TR (ms) | FOV (X, Y, Z) | Voxel size (mm) | Volumes | TE (ms) | TR (ms) | FOV (X, Y, Z) | Voxel size (mm) |
| Bonn | 30 | 2000 | 64 x 64 x 30 | 3 x 3 x 5 | 300 | 3.06 | 2300 | 240 x 256 x 176 | 1 x 1 x 1 |
| Erlangen | 34 | 3000 | 64 x 64 x 30 | 3 x 3 x 5 | 300 | 2.98 | 2300 | 240 x 256 x 176 | 1 x 1 x 1 |
| Göttingen | 30 | 2000 | 64 x 64 x 30 | 3 x 3 x 6 | 300 | 2.96 | 2300 | 256 x 256 x 176 | 1 x 1 x 1 |
| Homburg | 30 | 2000 | 64 x 64 x 30 | 3 x 3 x 5 | 300 | 2.98 | 2300 | 240 x 256 x 176 | 1 x 1 x 1 |
| Leipzig | 30 | 2000 | 64 x 64 x 30 | 3 x 3 x 5 | 300 | 2.98 | 2300 | 240 x 256 x 176 | 1 x 1 x 1 |
| München (TU) | 30 | 2000 | 64 x 64 x 30 | 3 x 3 x 5 | 300 | 2.98 | 2300 | 240 x 256 x 176 | 1 x 1 x 1 |
| Rostock | 30 | 2200 | 64 x 64 x 34 | 3.5 x 3.5 x 3.5 | 300 | 4.82 | 2500 | 256 x 256 x 192 | 1 x 1 x 1 |
| Tübingen | 30 | 2000 | 64 x 64 x 30 | 3 x 3 x 5 | 300 | 2.96 | 2300 | 240 x 256 x 176 | 1 x 1 x 1 |
| Ulm | 30 | 2000 | 64 x 64 x 30 | 3 x 3 x 5 | 300 | 2.05 | 2300 | 240 x 256 x 192 | 1 x 1 x 1 |

**rsfMRI – resting-state functional magnetic resonance imaging, MRI – magnetic resonance imaging, TE – echo time, TR – repetition time, FOV – field of view**

Bonn – University of Bonn, German Center for Neurodegenerative Diseases (DZNE), University Hospital Bonn
Erlangen – University Hospital Erlangen
Göttingen – Medical University Göttingen
Homburg – Saarland University Hospital
Leipzig – Max-Planck-Institute for Human Cognitive and Brain Sciences
TU München – Technical University of Munich
Rostock – University Hospital Rostock, German Center for Neurodegenerative Diseases (DZNE)
Tübingen – University Hospital Tübingen, Centre for Neurology, Hertie-Institute for Clinical Brain Research
Ulm – Ulm University

**Table S3** Detailed neurotransmitter map information incl. tracer, number of subjects, age, sex and study.

| Neurotransmitter | Tracer | N | Age | Sex ratio (% male subjects) | Study (doi) |
|---|---|---|---|---|---|
| 5-HT1a | way100635 | 35 | 26.3 ± 5.20 | 51.00 % | 10.1016/j.neuroimage.2012.07.001 |
| 5-HT1b | p943 | 23 | 28.7 ± 7.00 | 65.00 % | 10.1016/j.neuroimage.2012.07.001 |
| 5-HT2a | altanserin | 19 | 28.2 ± 5.70 | 58.00 % | 10.1016/j.neuroimage.2012.07.001 |
| 5-HTT | dasb | 18 | 30.5 ± 9.50 | 67.00 % | 10.1016/j.neuroimage.2012.07.001 |
| D1 | sch23390 | 13 | 33.0 ± 13.0 | 46.00 % | 10.1007/s00259-017-3645-0 |
| D2 | raclopride | 7 | 24.0 ± 2.00 | 100.00 % | 10.1038/jcbfm.2015.53 |
| DAT | fpcit | 174 | 61.0 ± 11.0 | 62.64 % | 10.1038/s41598-018-22444-0 |
| FDOPA | fluorodopa | 12 | 55.1 ± 16.6 | 50.00 % | 10.33588/imagendiagnostica.901.2 |
| GABAa | flumazenil | 6 | 43.0 ± 4.00 | 100.00 % | 10.1038/s41598-018-22444-0<br>10.1038/jcbfm.2011.177 (original study) |
| MU | carfentanil | 204 | 32.3 ± 10.8 | 64.71 % | 10.1038/mp.2017.183 |
| NET | mrb | 10 | 33.3 ± 10.0 | 60.00 % | 10.1007/s00259-016-3590-3 |

**Table S4** Contrast peak voxels (HC>bvFTD) incl. MNI coordinates, corresponding anatomical region, t-value and cluster size for fALFF and GMV.

| | **MNI coordinates at peak voxel (mm)** | | | **Corresponding anatomical region** | **T (peak-level)*** | **Cluster size (voxels)** |
|---|---|---|---|---|---|---|
| | **x** | **y** | **z** | | | |
| fALFF | -42 | 6 | 18 | Left inferior frontal gyrus | 6.76 | 6973 |
| | 39 | -72 | 45 | Right inferior lateral parietal lobe | 5.64 | 927 |
| | -39 | -27 | 60 | Left postcentral gyrus | 5.17 | 310 |
| | -54 | -18 | -6 | Left superior temporal gyrus | 4.39 | 113 |
| | -18 | 3 | 24 | Left caudate nucleus | 4.34 | 70 |
| | 9 | 6 | 0 | Right lateral temporal ventricle | 4.91 | 45 |
| GMV | -45 | -24 | 12 | Left superior temporal gyrus | 7.52 | 20308 |

* cluster-level threshold: p-uncorrected = .001 with permuted cluster threshold (fALFF: 42 voxels, GMV: 38 voxels)

**fALFF – fractional amplitude of low frequency fluctuations, GMV – gray matter volume**

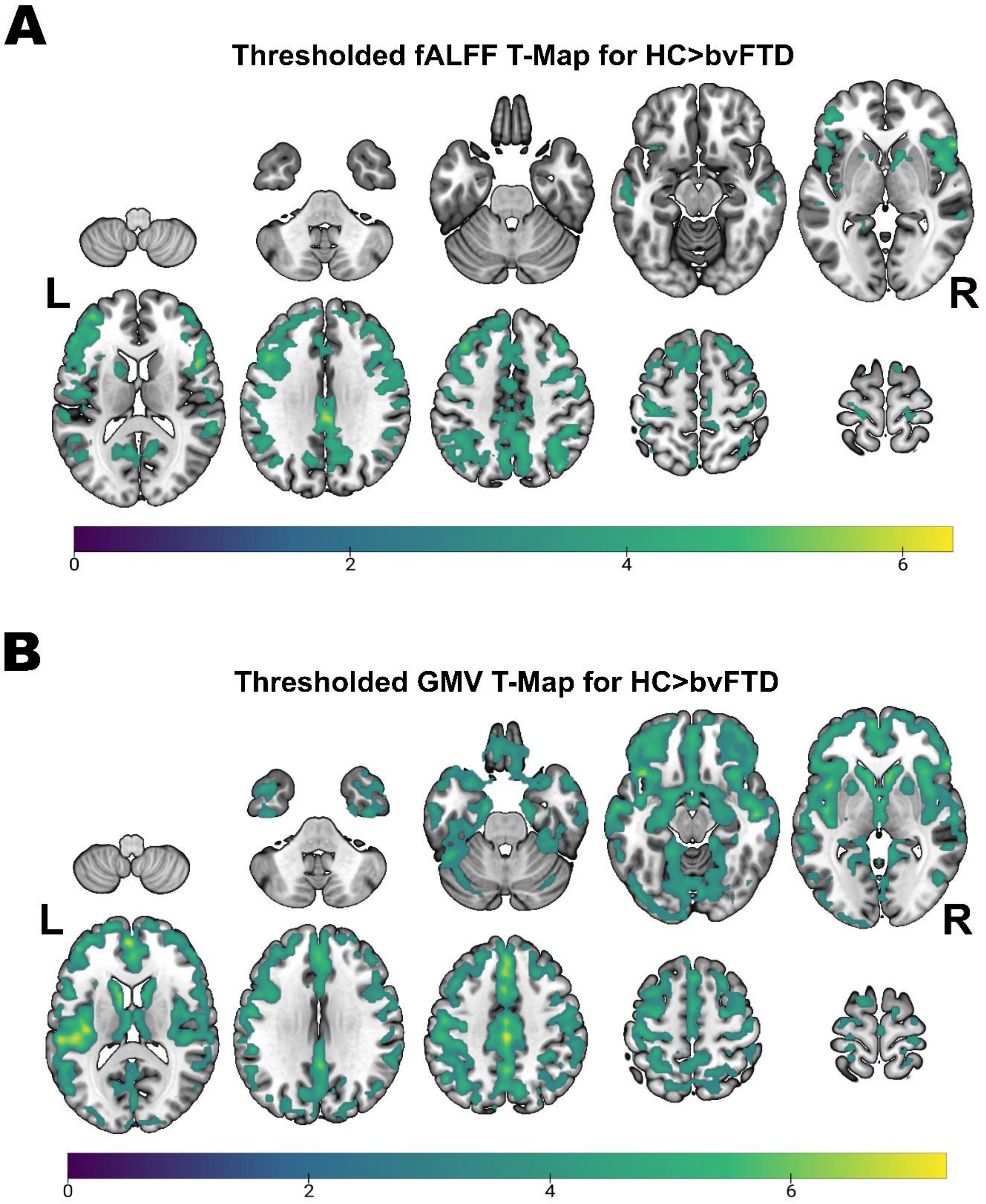


**Fig. S1 Detailed voxel-wise results for fALFF and GMV group comparisons.** Thresholded fALFF t-map **(A)** and thresholded GMV t-map **(B)** for HC>bvFTD using a permutation-based threshold (1000 permutations permuting group labels) at cluster-level p<.05 and voxel-level p<.001.

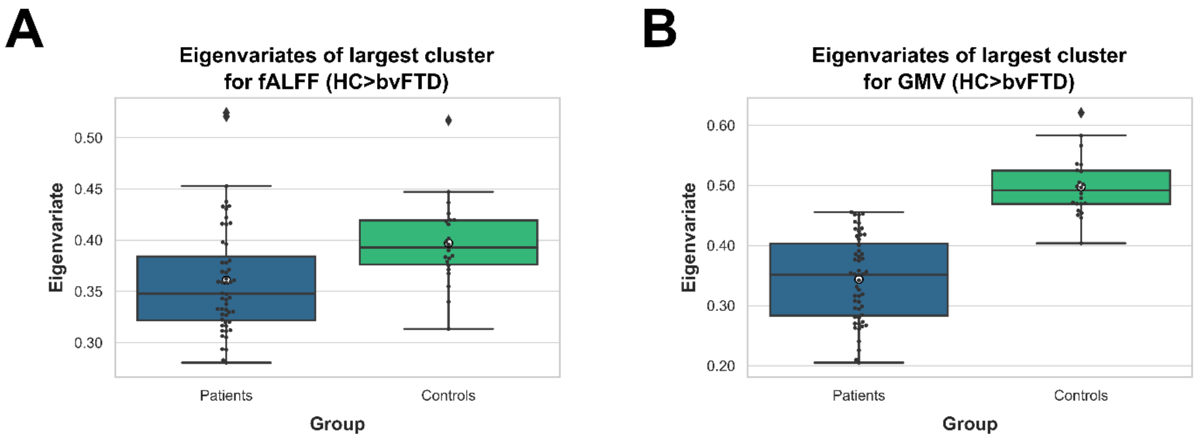


**Fig. S2 Eigenvariates from fALFF and GMV for bvFTD patients and controls.** Eigenvariates were derived from the largest cluster in the HC>FTD contrasts for fALFF (A) and GMV (B) ($N_{bvFTD}$ = 52, $N_{HC}$ = 22). Means are represented by white circles.

**Table S5** Fisher's z transformed Spearman correlations of neurotransmitter maps with fALFF and gray matter volume.

| **Neurotransmitter** | | **fALFF** | **GMV** |
|---|---|---|---|
| 5-HT1a | r (Spearman) | -.0316 | -.0787 |
| | p | .5531 | .1447 |
| 5-HT1b | r (Spearman) | -.2152* | -.1143 |
| | p | <.0001 | .0636 |
| 5-HT2a | r (Spearman) | -.1653* | -.0771 |
| | p | .0015 | .2340 |
| 5-HTT | r (Spearman) | -.0219 | -.0525 |
| | p | .7401 | .4630 |
| D1 | r (Spearman) | -.0613 | -.0660 |
| | p | .1620 | .3018 |
| D2 | r (Spearman) | -.1772* | -.0410 |
| | p | .0057 | .4129 |
| DAT | r (Spearman) | .0004 | -.0171 |
| | p | .9963 | .8054 |
| FDOPA | r (Spearman) | -.0225 | -.0144 |
| | p | .7360 | .8351 |
| GABAa | r (Spearman) | -.1256* | -.0589 |
| | p | .0157 | .3807 |
| MU | r (Spearman) | -.0676 | -.1104 |
| | p | .2733 | .1820 |
| NET | r (Spearman) | -.1353* | -.0116 |
| | p | .0143 | .8188 |
| $p_{FDR}$ | | p = .0157 | - - - |

* FDR-corrected significant correlation (p<.05)

**fALFF – fractional amplitude of low frequency fluctuations, GMV – gray matter volume**

**Table S6** Fisher's z transformed Spearman correlations between significant fALFF-neurotransmitter correlations and neuropsychological test data.

| Neurotransmitter | | VF | BNT | MMSE | TMTB | AES | FrSBe Freq | FrSBe Dist | FrSBe Dist EF | CDR-FTLD | $p_{FDR}$ |
|---|---|---|---|---|---|---|---|---|---|---|---|
| 5-HT1b | r (Spearman) | .0711 | .0260 | .0633 | -.0638 | .0360 | -.0538 | .1493 | .1168 | -.2186 | --- |
| | p | .6275 | .8592 | .6625 | .7115 | .8372 | .7623 | .4395 | .5106 | .1491 | |
| 5-HT2a | r (Spearman) | .0405 | .0540 | .0479 | -.0885 | -.0126 | .0876 | .1787 | .0782 | -.1077 | --- |
| | p | .7824 | .7127 | .7411 | .6078 | .9426 | .6221 | .3537 | .6601 | .4812 | |
| D2 | r (Spearman) | .3650* | .2159 | .4021* | -.1688 | .1493 | .2685 | .4489 | .4224 | -.1248 | p = .0146 |
| | p | .0099 | .1363 | .0038 | .3250 | .3919 | .1248 | .0146 | .0128 | .4142 | |
| GABAa | r (Spearman) | .0369 | -.0341 | .0793 | -.1208 | .0374 | .1562 | .1624 | .0761 | -.1303 | --- |
| | p | .8012 | .8162 | .5841 | .4828 | .8309 | .3778 | .4000 | .6689 | .3936 | |
| NAT | r (Spearman) | .3719 | .2657 | .3797 | -.1635 | .1437 | .2880 | .3238 | .3521 | -.0838 | p = .0085 |
| | p | .0085 | .0650 | .0065 | .3407 | .4102 | .0986 | .0866 | .0411 | .5841 | |

* FDR-corrected significant correlation (p<.05)

**fALFF – fractional amplitude of low frequency fluctuations, VF – Verbal Fluency, BNT – Boston Naming Test, MMSE – Mini Mental State Exam, TMTB – Trail Making Test B, AES – Apathy Evaluation Scale, FrSBe Freq – Frontal Systems Behavior Scale Frequency Score, FrSBe Dist – Frontal Systems Behavior Scale Distress Score, FrSBe Dist EF – Frontal Systems Behavior Scale Distress Score for Executive Functioning subscale, CDR-FTLD – Clinical Dementia Rating Frontotemporal Lobar Degeneration**

**Table S7** Spearman correlations between fALFF Eigenvariates from largest cluster (HC>bvFTD) and neuropsychological test data.

| **Eigenvariates correlation with** | **r (Spearman's rho)** | **p** |
|---|---|---|
| VF | .2459 | .0885 |
| BNT | .2076 | .1524 |
| MMSE | .2629 | .0651 |
| TMT-B | -.3188 | .0581 |
| AES | .0000 | 1.0000 |
| FrSBe Freq | -.0519 | .7709 |
| FrSBe Dist | .0022 | .9909 |
| FrSBe Dist EF | -.0543 | .7602 |
| CDR-FTLD | -.3385 | .0229 |
| $p_{FDR}$ | --- | |

**VF – Verbal Fluency, BNT – Boston Naming Test, MMSE – Mini Mental State Exam, TMTB – Trail Making Test B, AES – Apathy Evaluation Scale, FrSBe Freq – Frontal Systems Behavior Scale Frequency Score, FrSBe Dist – Frontal Systems Behavior Scale Distress Score, FrSBe Dist EF – Frontal Systems Behavior Scale Distress Score for Executive Functioning subscale, CDR-FTLD – Clinical Dementia Rating Frontotemporal Lobar Degeneration, FDR – False Discovery Rate**

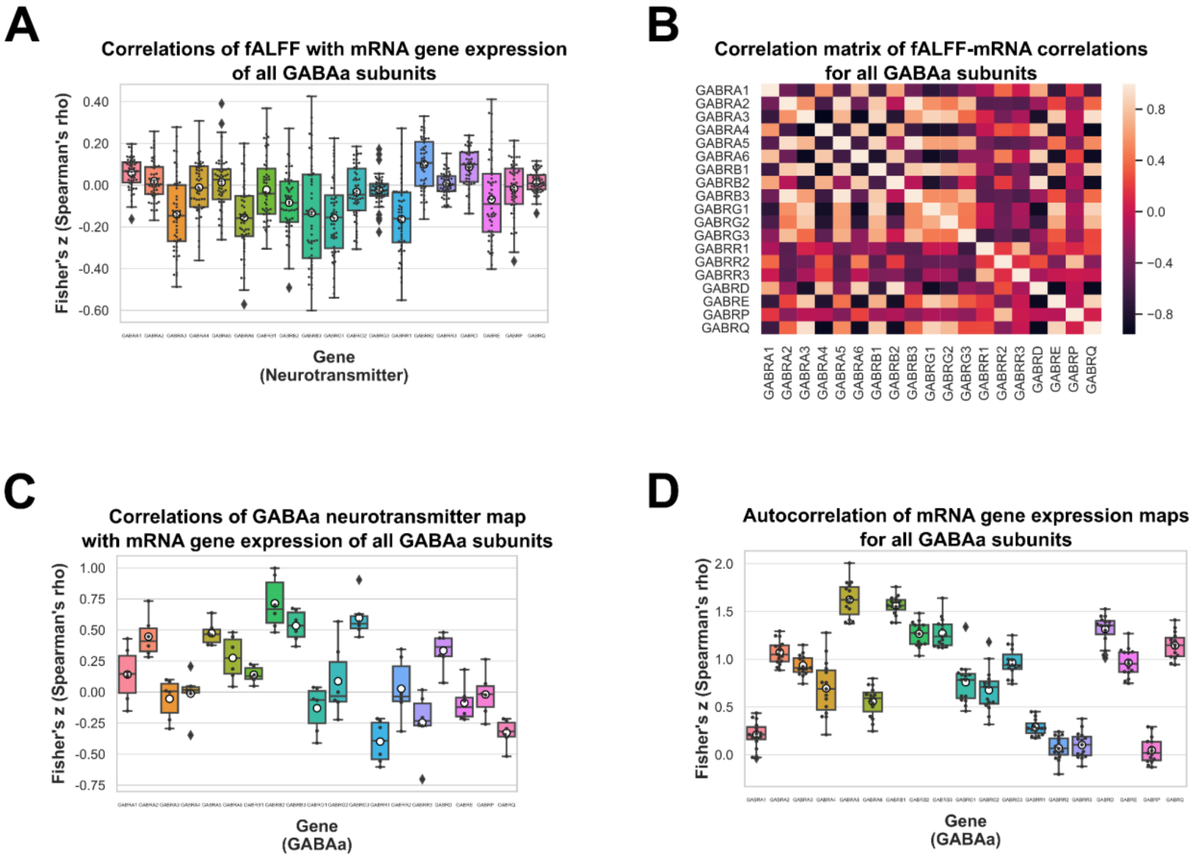


**Fig. S3 Results for mRNA gene expression maps of all GABAa subunits.** Correlations of mRNA gene expression maps with fALFF for all GABAa subunits (N = 52) (A) and their corresponding correlation matrix (B), their correlation with the GABAa neurotransmitter map ($N_{donors}$ = 6) (C), and their mRNA autocorrelations ($N_{donors}$ = 6) (D). The genes encoding the nineteen GABAa subunits include GABRA1-6, GABRB1-3, GABRG1-3, GABRR1-3, GABRD, GABRE, GABRP and GABRQ. Means are represented by white circles.
